# Analytical and Clinical Validation of an Amplicon-based Next Generation Sequencing Assay for Ultrasensitive Detection of Circulating Tumor DNA

**DOI:** 10.1101/2021.08.03.21261575

**Authors:** Jonathan Poh, Kao Chin Ngeow, Michelle Pek, Kian-Hin Tan, Jing Shan Lim, Hao Chen, Choon Kiat Ong, Jing Quan Lim, Soon Thye Lim, Chwee Ming Lim, Boon Cher Goh, Yukti Choudhury

## Abstract

Next-generation sequencing of circulating tumor DNA presents a promising approach to cancer diagnostics, complementing conventional tissue-based diagnostic testing by enabling minimally invasive serial testing and broad genomic coverage through a simple blood draw to maximize therapeutic benefit to patients. LiquidHALLMARK® is an amplicon-based next-generation sequencing assay developed for the genomic profiling of plasma-derived cell-free DNA. The comprehensive 80-gene panel profiles point mutations, insertions/deletions, copy number alterations, and gene fusions, and further detects oncogenic viruses (EBV and HBV) and microsatellite instability. Here, the analytical and clinical validation of the assay is reported. Analytical validation using reference genetic materials demonstrated a sensitivity of 99.38% for point mutations and 95.83% for insertions/deletions at 0.1% variant allele frequency (VAF), and a sensitivity of 91.67% for gene fusions at 0.5% VAF, with high specificity even at 0.1% VAF (99.11% per-base). The limit of detection for copy number alterations, EBV, HBV, and microsatellite instability were also empirically determined. Orthogonal comparison of *EGFR* variant calls made by LiquidHALLMARK and a reference allele-specific PCR method for 355 lung cancer specimens revealed an overall concordance of 93.80%, while external validation with cobas® EGFR Mutation Test v2 for 50 lung cancer specimens demonstrated an overall concordance of 84.00%, with a 100% concordance rate for *EGFR* variants above 0.4% VAF. Clinical application of LiquidHALLMARK in 1,592 consecutive patients demonstrated a high detection rate (74.8% alteration-positive in cancer samples) and broad actionability (50.0% of cancer samples harboring alterations with biological evidence for actionability). Among ctDNA-positive lung cancers, 72.5% harbored at least one biomarker with a guideline-approved drug indication. These results establish the high sensitivity, specificity, accuracy, and precision of the LiquidHALLMARK assay and supports its clinical application for blood-based genomic testing.

## Introduction

The diagnosis and subclassification of cancers, fundamental to any cancer treatment algorithm, has historically been achieved through the study of the primary tumor tissue. The availability of high-quality diagnostic material suitable for high-end genomic technologies, including multiplexed tissue-based genotyping, is often limiting and associated with high risks and costs [1]. Indeed, for diseases such as advanced-stage, non-squamous, non-small-cell lung cancers (NSCLCs), approximately 30% of tumor samples yield insufficient or inadequate tissue for successful molecular genotyping [2,3]. Such limitations have positioned minimally invasive ‘liquid biopsy’-based diagnostics as an attractive alternative for cancer management. Profiling of cell-free DNA (cfDNA) in blood to detect circulating tumor DNA (ctDNA) [4,5] has tremendous potential in overcoming the limits of sampling frequency and tumor accessibility, as well as the requirement of clinically overt disease imposed by traditional tumor biopsies. Such advances are reflected in clinical practice upgrades, such as the National Comprehensive Cancer Network (NCCN) guidelines for NSCLC now encouraging the use of liquid biopsies in cases where tissue biopsies may not be feasible [6], particularly as many patients harbor biomarkers that can be targeted by current or emerging therapies [7]. Precisely due to its non-invasive nature, the utility of cfDNA profiling extends to every stage of clinical management, from diagnosis, selection of targeted treatments, minimal residual disease detection to identifying acquired drug resistance, in the background of spatial and temporal tumor heterogeneity [8,9]. Recent years have seen a number of approvals for cfDNA-based tests in the companion diagnostic setting based on polymerase chain reaction (PCR), such as *EGFR* in the cobas® EGFR Mutation Test v2 and *PIK3CA* in the therascreen® PIK3CA RGQ PCR Kit [10], and very recently based on next-generation sequencing (NGS) [11,12].

Broad molecular coverage offered by NGS-based panel tests for ctDNA profiling allows the evaluation of multiple markers from the same plasma cfDNA sample, improving the detection of ctDNA in a sample below the technical lower limit of detection of the assay. Despite this, alterations identified through routine analysis of tumor tissue are detected in comprehensive ctDNA analysis with a sensitivity of 80 – 90% [13–16]. Successful ctDNA detection is known to be affected by the anatomical site of disease, tumor burden, and the sensitivity/specificity profile of the assay [9]. For utility in multiple clinical settings, cfDNA-based tests should ideally be able to detect ctDNA fractions <1%, against the setting of inherent error rates of NGS platforms of 0.1 – 1% [17]. The technical limits of detection of ctDNA platforms have been tremendously improved by ultra-deep sequencing of targeted variants, error-suppression methods, and statistical error-modeling [18,19]. Still, the challenges of detecting low-level ctDNA accurately, particularly for low disease burden (correlated to overall ctDNA amount) hamper concordance between tissue and cfDNA tests, leading to significant discordances between multiple cfDNA platforms at <1% ctDNA levels [20], and limit the generalizability of any one ctDNA biomarker. Negative ctDNA results are typically reflexed to tissue genotyping for NSCLC [21] and testing for *PIK3CA* in hormone-positive breast cancers [22].

Both biological and technical factors work in concert to determine the ability to detect ctDNA accurately, and overcoming technical limitations presents near-term opportunities to enhance this ability. Two basic approaches to broadly capture cfDNA are amplicon (PCR)-based and hybridization-capture based methods [23]. Broad cfDNA panel tests that have been validated are presently largely based on the latter approach [15,24], with amplicon-based assays being relatively limited in panel breadth [25]. In principle, the amplicon-based target capture utilizing a pair of primers per target site, as opposed to a single hybridization probe, affords a greater degree of specificity for the enrichment of desired target regions. In turn, this enables interrogation of limiting amounts of ctDNA (as in low tumor burden samples), and enriches the density of informative sequencing reads which are on-target. Analytical sensitivities of amplicon-based methods are reportedly superior [25,26] compared to hybridization-based methods [15,24] for single nucleotide variants (SNVs) and insertion/deletion events (INDELs), in the biologically relevant sub-1% ctDNA range. While multiple analytical and post-analytical factors differ between individual methods, including variant calling algorithms, better sensitivity can at least be partially attributed to the superior on-target capture rate of amplicon-based enrichment methods. At present, there is a paucity of comprehensive validation studies for NGS-based cfDNA tests, particularly those based on amplicon-based enrichment, which possess technical advantages in terms of simpler workflow, effectiveness with limited input DNA, and higher on-target capture rates.

In this work, we present the analytical validation of an amplicon-based NGS assay, LiquidHALLMARK®, with broad target coverage for clinically actionable genes. The assay incorporates two degrees of error-correction, through the use of unique molecular identifiers (UMI) introduced at target capture, and position-specific background error suppression through statistical modeling of noise inherent to samples and their processing. In addition to the profiling of SNVs, INDELs, copy number alterations (CNAs), and gene fusions (known and novel), the assay is the first NGS-based cfDNA panel test to incorporate the detection of cancer-causing viruses, including Epstein-Barr Virus (EBV) and Hepatitis B virus (HBV), and detection of microsatellite instability (MSI) in cfDNA, validated through multiple orthogonal comparisons. We demonstrate the clinical applicability of LiquidHALLMARK by quantitatively presenting the diagnostic findings in consecutive samples received for routine clinical testing at our Clinical Laboratory Improvement Amendment (CLIA)-certified, College of American Pathologists (CAP)-accredited laboratory. LiquidHALLMARK is applicable for ultra-sensitive detection of ctDNA in multiple cancer types, with a limit of detection of 0.1% VAF for SNVs and INDELs, with which clinically actionable alterations can be identified and monitored.

## Results

### Assay design/specifications

LiquidHALLMARK is a targeted NGS-based sequencing diagnostic assay that profiles 80 cancer-related genes using cfDNA isolated from stabilized peripheral whole blood-derived plasma of cancer patients (**Fig 1**). The assay detects SNVs and INDELs in 77 genes, with nearly full exonic coverage in five genes and carefully curated inclusion of hotspot and critical regions in select oncogenes and tumor suppressors, spanning 58.8 kb of genome coverage. In addition, CNAs in 18 genes, and structural rearrangements in 10 genes, including the detection of both novel and pre-determined gene fusion events, are captured by the assay. Further, the assay detects MSI, as well as the presence of HBV and EBV, which are key oncogenic signatures in colorectal cancer, nasopharyngeal cancer, and liver cancer, respectively. Although less comprehensive than conventional 500-gene tissue-based NGS panels, rational inclusion of key cancer-specific and pan-cancer driver genes, non-genetic biomarkers, as well as targets for which FDA-approved drugs, investigational drugs, or clinical trials are available (**Supplementary Fig 1**), has provided a diagnostic yield of 74.8% in all cancer samples processed to date (n = 1521).

**Figure 1.**
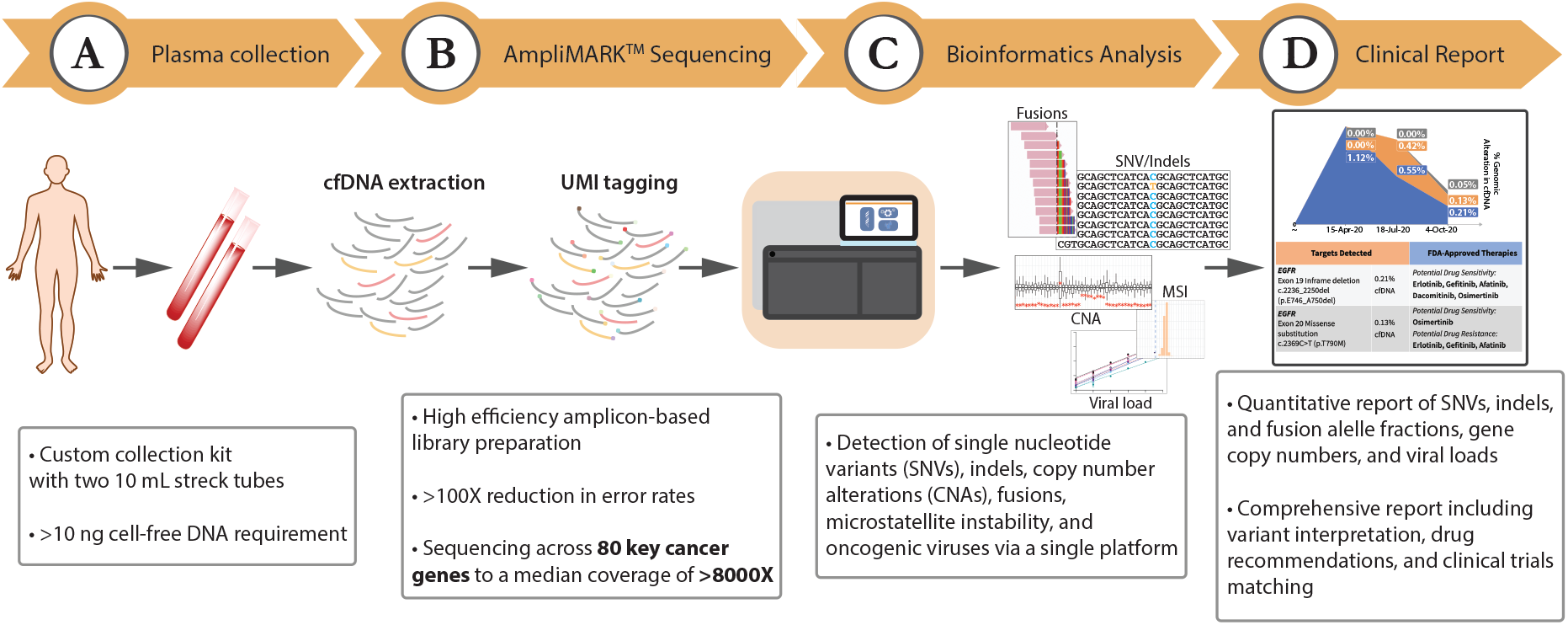
Overview of the LiquidHALLMARK^®^ assay workflow. **(A)** Stabilized whole blood is collected using custom collection kits and cell-free DNA (cfDNA) isolated from plasma. Up to 20 ng of cfDNA is used for library preparation. **(B)** Targeted genomic regions are tagged with oligonucleotide barcodes and enriched via high efficiency amplicon-based library preparation before being sequenced to a median coverage of >8000X. UMI, unique molecular identifier. **(C)** Sequencing data is bioinformatically deconvoluted, with molecular barcodes enabling error suppression rates above 100X. **(D)** Somatic variants and other biomarkers are quantitative reported, with comprehensive interpretation encompassing drug actionability and clinical trial eligibility.

LiquidHALLMARK utilizes a highly optimized amplicon-based library preparation method combined with UMI labelling to efficiently recover unique molecules from just 20 ng input cfDNA, sequencing to a median unique coverage of 8354X (range 42.0 – 98.4% recovery of unique DNA molecules). With a median amplicon length of 166 bp, which approximates the median length of cfDNA [27], the assay generates highly uniform sequencing profiles; on average 97.1% of the panel has a depth no less than 0.2X the mean panel coverage and 79.0% of the panel has a depth no less than half the mean panel coverage. In addition, UMI sequencing combined with statistical background error-modelling approaches enable error suppression of more than 100X, allowing the confident calling of SNVs/INDELs above noise levels at as low as 0.01% variant allele frequency (VAF; **Supplementary Fig 2**).

### SNV/INDEL detection performance

To determine the analytical sensitivity, precision, and accuracy of the LiquidHALLMARK assay in detecting SNVs and INDELs, the reference standard sets HD780 (**Fig 2A**) and diluted Tru-Q (**Fig 2B**; Horizon Discovery), comprising a total of 39 unique clinically relevant SNVs and three INDELs in 19 genes across 5% (3.6 – 6.5%), 1% (1.0 – 1.3%), and 0.1% (0.10 – 0.13%) VAF verified by droplet digital PCR (ddPCR), were run in triplicates across multiple operators, two reagent lots, and two NextSeq 550 sequencers. In addition, the HD786 reference standard (Horizon Discovery) comprising six SNVs and four INDELs at 5% (5.0 – 5.6%) and one INDEL at 2.5% was run by two different operators. Across a total of 674 SNV/INDEL observations, only one INDEL (EGFR E746_A750del) and three SNVs (KRAS G12D, Q61H, Q61L) were missed at 0.1% VAF, and one SNV at 1% VAF (KRAS Q61H). Because the KRAS Q61 locus consistently gave a depth <6% of the mean panel coverage (average depth at locus <850X) specifically in the Tru-Q standards, the KRAS Q61H and KRAS Q61L variants covered by this locus were excluded from all sensitivity analyses. Hence, these results yield a sensitivity of 100% at 5% and 1% VAF, and 98.92% at 0.1% VAF, with individual sensitivities of 99.38% and 95.83% for SNVs and INDELs respectively (**Table 1**). The per-base analytical specificity prior to noise filtering was determined to be 99.98% at 5% and 1% VAF, and 99.11% at 0.1% VAF using a total interrogable panel size of 58.8 kb, giving a false positive rate of 0.022% at 5% VAF, 0.025% at 1% VAF, and 0.90% at 0.1% VAF. These results confirm a 90% limit of detection (LoD_90_) of 0.1% VAF for both SNVs and INDELs. Across the HD780 reference standards, pairwise analysis gave an overall reproducibility of 100% at 5% and 1% VAF, and 97.22% at 0.1% VAF, with no noticeable difference within or between operators, reagent lots, and sequencers (**Fig 2A**). Quantitative accuracy of variant calls was high, with a good degree of correlation between observed and expected VAFs (**Fig 2C**; R^2^ = 0.97, y = 0.90).

**Figure 2.**
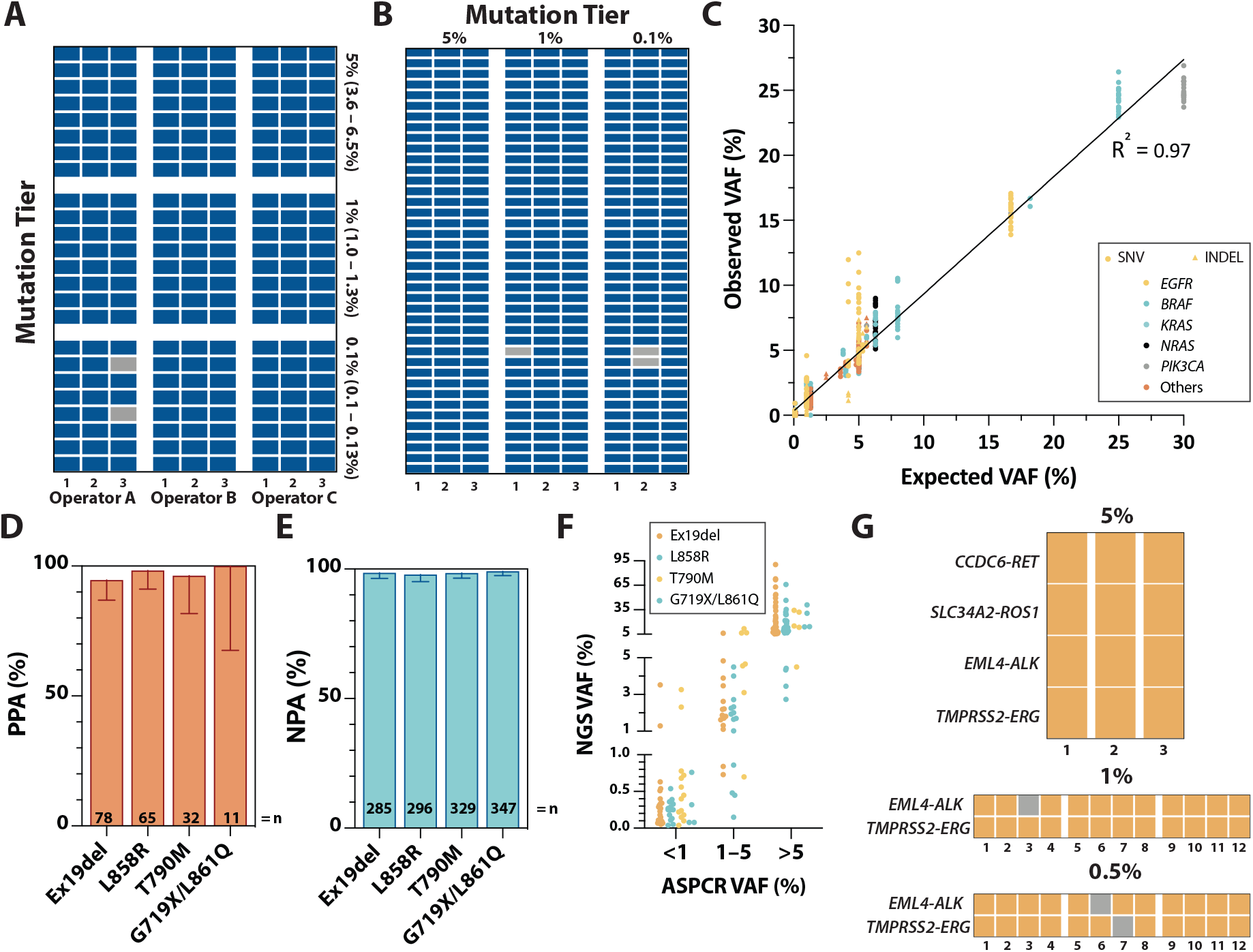
Analytical performance for (A-F) SNV/INDEL and (G) fusion detection. Hit/miss observations across different SNV/INDEL mutation tiers and operators/repeats in **(A)** HD780 and **(B)** Tru-Q reference standard materials. Each row represents a unique SNV/INDEL variant. Detected SNV/INDEL variants are highlighted in blue and missed variants in gray. **(C)** Correlation between expected and observed SNV/INDEL variant allele frequencies (VAF) based on linear regression. Circles represent SNVs and triangles INDELs. Each color represents a unique gene. (**D**) Positive percent agreement (PPA), (**E**) negative percent agreement (NPA) and (**F**) positive VAF concordance of LiquidHALLMARK *EGFR* variant calls relative to allele-specific PCR (AS-PCR) in 355 clinical lung cancer samples. Error bars in **D** and **E** represent 95% CI. Each color in (**F**) represents a unique class of EGFR variant; ex19del, exon 19 deletion. (**G)** Hit/miss observations across different fusion mutation tiers and repeats in HD786 reference standard and contrived admixtures of fragmented cell-line DNA. Each row represents a unique fusion. Detected SNV/INDEL variants are highlighted in orange and missed variants in gray.

**Table 1.** Analytical sensitivity of the LiquidHALLMARK assay.

| Mutation Tier | Variant Type | Sensitivity (95% CI) |
| --- | --- | --- |
| 3.6 – 6.5% | SNVs | 100% (97.91%, 100%) |
|  | INDELs | 100% (89.28%, 100%) |
|  | Fusions | 100% (77.19%, 100%) |
| 1.0 – 1.3% | SNVs | 100% (97.68%, 100%) |
|  | INDELs | 100% (86.20%, 100%) |
|  | Fusions | 95.45% (78.20%, 99.77%) |
| 0.5% | Fusions | 91.67% (74.15%, 98.52%) |
| 0.1 – 0.13% | SNVs | 99.38% (96.59%, 99.97%) |
|  | INDELs | 95.83% (79.76%, 99.79%) |

To validate the clinical accuracy of LiquidHALLMARK for SNV and INDEL variant calls, 355 lung cancer specimens received for clinical testing were concurrently analyzed for 10 specific *EGFR* mutations, including exon 19 deletions, L858R, T790M, G719X, and L861Q, using a reference allele-specific PCR (AS-PCR) method [28]. LiquidHALLMARK and AS-PCR were concordant for 127 positive calls (positive percent agreement, PPA 95.49%; 95% CI, 90.51 – 97.92%; NGS VAF 0.03 – 90.43%) and 206 negative calls (negative percent agreement, NPA 92.79%; 95% CI, 88.61 – 95.52%), giving an overall concordance of 93.80% (95% CI, 90.80 – 95.87%; **Table 2**). When segregated by *EGFR* variant type (exon 19 deletion, L858R, T790M, or G719X/L861Q), both PPA and NPA remained high (>94% and >97% respectively) for all four *EGFR* variant types (**Fig 2D-E**), with good agreement between quantitative calls made by both methods (**Fig 2F**). In total, four exon 19 deletion, one L858R, and one T790M variants were missed by LiquidHALLMARK at detected VAFs close to or below the LoD_90_ (AS-PCR VAF 0.03 – 0.16%). LiquidHALLMARK detected three additional G719X, four exon 19 deletion, five L858R, and five T790M variants (NGS VAF 0.02 – 0.50%); 10 of these were detected specifically by AS-PCR melt curve analysis below the reportable range. Of the remaining seven discordant variants, six were detected close to or below the established LoD of AS-PCR, with one discordant exon 19 deletion (L747_P753delinsS) detected at 0.5% VAF in LiquidHALLMARK. These findings corroborate the high sensitivity and accuracy of the LiquidHALLMARK assay.

**Table 2.**
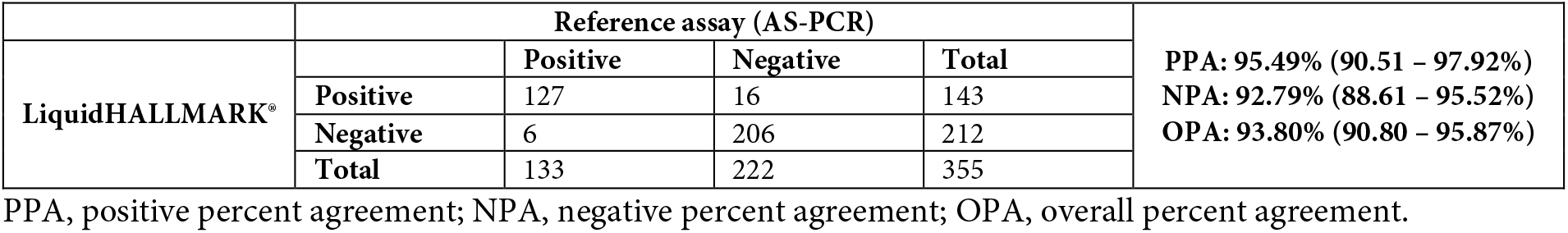
Comparison of LiquidHALLMARK with a reference AS-PR assay for the detection of EGFR L858R, exon 19 deletion, T790M, and uncommon G719X/L861Q variants.

| LiquidHALLMARK® | Reference assay (AS-PCR) |  |  |  | PPA: 95.49% (90.51 – 97.92%)<br>NPA: 92.79% (88.61 – 95.52%)<br>OPA: 93.80% (90.80 – 95.87%) |
| --- | --- | --- | --- | --- | --- |
|  |  | Positive | Negative | Total |  |
|  | Positive | 127 | 16 | 143 |  |
|  | Negative | 6 | 206 | 212 |  |
|  | Total | 133 | 222 | 355 |  |

PPA, positive percent agreement; NPA, negative percent agreement; OPA, overall percent agreement.

### Fusion detection performance

To determine the analytical sensitivity of the assay for the detection of gene fusions, the HD786 reference standard (Horizon Discovery) and contrived admixtures of fragmented fusion positive cell-line DNA were used to probe four gene fusions (*EML4-ALK, SLC34A2-ROS1, TMPRSS2-ERG*, and *CCDC6-RET*) across 5%, 1%, and 0.5% VAF. A total of 12 fusions were tested at 5% VAF across three replicates and 24 fusions at 1% and 0.5% VAF across 12 replicates each, with 1 *EML4-ALK* fusion missed at 1% VAF and 1 *EML4-ALK* fusion and 1 *TMPRSS2-ERG* fusion missed at 0.5% VAF (**Fig 2G**), giving a sensitivity of 100%, 95.45%, and 91.67% respectively (**Table 1**). Thus, the LoD_90_ was empirically determined to be 0.5% VAF for gene fusions.

Due to a lack of fusion-positive genetic material, the platform technology was further validated for the ability to detect novel fusions by analyzing 29 NK/T-lymphoma tissue samples for PD-L1 3’-untranslated region (3’-UTR) rearrangement positivity [29]. Of these, nine samples tested positive for PD-L1 3’-UTR rearrangement and 20 samples tested negative; all calls were orthogonally confirmed by whole-genome, targeted, or Sanger sequencing, giving an overall concordance of 100% (**Supplementary Table 1**). In one sample where matched plasma was available, cfDNA testing identified the same PD-L1 3’-UTR rearrangement.

### Copy number alteration (CNA) detection performance

Validation of CNA detection performance was conducted using contrived admixtures of fragmented cell-line DNA from cell-lines well-established to harbor gene amplifications in *EGFR, ERBB2* (HER2), *NRAS*, and *CDK6* [30–32], as well as the HD786 reference standard which harbors focal amplification in *MET* and *MYC* at 4.5 and 9.5 copies respectively, instead of normal diploid two copies. Across 19 observations from 2.4 to 65.2 gene copies in the dilution series, all amplifications were positively detected, with a high degree of correlation between expected and observed copy numbers (R^2^ = 0.92, y = 0.70; **Fig 3A**). Based on gene amplifications confidently detectable above the range of normalized copy numbers determined from a set of selected reference plasma cfDNA samples (n = 27), the LoD for gene amplifications was determined to be a 1.5X increase from the normal diploid number, which is equivalent to a low-level amplification (copy number of 6) at 25% tumor fraction (TF) or a high-level amplification (copy number of 10) at 12.5% TF.

**Figure 3.**
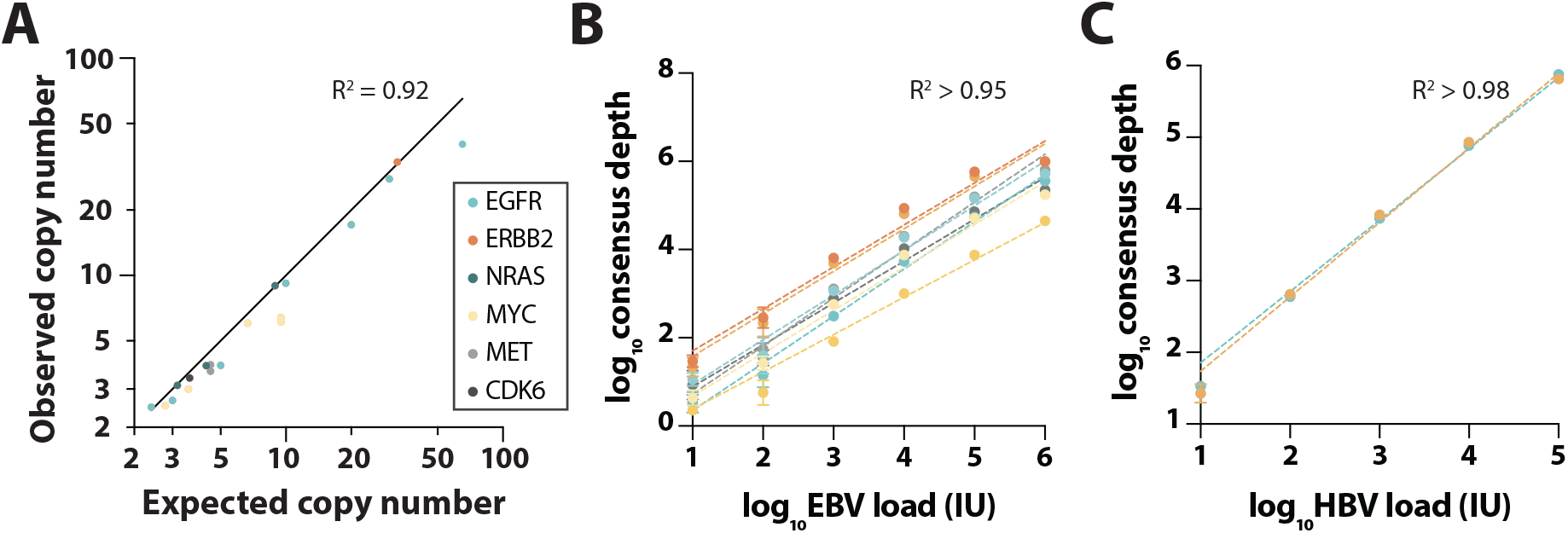
Validation of LiquidHALLMARK^®^ read count-based biomarkers. **(A)** Positive concordance of observed and expected copy numbers for amplification in six genes using HD786 reference standard and contrived admixtures of fragmented cell-line DNA. Observed consensus depth relative to **(B)** EBV and **(C)** HBV viral loads in plasma. Data is represented as mean ± SD from four replicate experiments. Each color represents a unique virus-specific amplicon. Correlation between consensus depth and viral load is based on linear regression.

To assess the concordance between gene amplification calls in plasma cfDNA and fluorescence *in situ* hybridization (FISH) in tissue, nine matched plasma cfDNA samples from *ERBB2* (HER2) amplification-positive samples were analyzed by LiquidHALLMARK. Six samples were positive for *ERBB2* (HER2) amplification in plasma cfDNA, giving a positive concordance between cfDNA and tissue testing of 66.67% (**Supplementary Table 2**).

### Detection performance for other biomarkers

In addition to detecting conventional biomarkers in SNVs, INDELs, gene fusions, and CNAs, the LiquidHALLMARK assay detects and quantifies viral loads of EBV and HBV in plasma cfDNA via the inclusion of amplicons specific for these viruses. EBV detection performance was verified by spiking plasma cfDNA with known viral loads of the external reference standard for EBV (1^st^ WHO International Standard for Epstein-Barr Virus, NIBSC code: 09/260) spanning six orders of magnitude. Based on the dilution series performed in four replicates, a high degree of linearity could be observed for all eight EBV-specific amplicons (R^2^ >0.95 for all amplicons, **Fig 3B**). These experiments also establish an LoD of 10 IU EBV within a sequencing run, equivalent to 2 IU/mL plasma when the routine 5 mL of plasma is used for cfDNA extraction. To validate the clinical performance of the assay for EBV detection, 19 clinical cfDNA specimens were analyzed by LiquidHALLMARK and an orthogonal PCR method [33]. A total of 16 positive calls and 1 negative call were observed by both methods, giving a PPA of 94.12% and an NPA of 50.00% (**Supplementary Table 3**).

HBV detection performance was assessed by spiking plasma cfDNA with known viral loads of the external reference standard for HBV (4th WHO International Standard for HBV DNA for NAT, NIBSC code: 10/266) spanning six orders of magnitude. Based on the dilution series performed in four replicates, a high degree of linearity of response was observed across both HBV-specific amplicons (R^2^ >0.98 for both amplicons, **Fig 3C**).

The detection threshold for microsatellite instability (MSI) was determined using 91 plasma cfDNA samples and 29 FFPE tissue samples with no known MSI, with the threshold for calling MSI set as the 95^th^ percentile of deletion length (for each of the six MSI loci) detected across all microsatellite stable (MSI-negative) samples. The LoD for MSI was validated using dilutions of DNA from the MSI-positive cell-line RKO in a wild-type cfDNA background and confirmed to be 5% tumor fraction (**Supplementary Fig 3**). To compare concordance for MSI detection between plasma cfDNA and tissue samples, matched cfDNA was analyzed for 36 clinical samples in which microsatellite status was determined in tumor tissue by fragment size analysis. LiquidHALLMARK and tissue testing were concordant for 4 positive and 31 negative calls, giving an overall concordance of 97.22% (PPA 80.00%, NPA 100%; **Supplementary Table 2**).

### Clinical validation for detection of *EGFR* alterations

To assess the clinical accuracy of LiquidHALLMARK for *EGFR* SNV and INDEL variant calls, LiquidHALLMARK was externally validated against the FDA-approved cobas EGFR Mutation Test v2. Fifty samples analyzed by LiquidHALLMARK, including 38 positive and 12 negative for *EGFR* variants covered by the cobas assay were submitted to the Mayo Clinic Laboratories (Rochester Main Campus, MN) for cobas testing to benchmark the concordance between LiquidHALLMARK and cobas calls. Of the 50 samples, LiquidHALLMARK and cobas were concordant for 31 positive (PPA 96.88%; 95% CI, 84.26 – 99.84%) and 11 negative variant calls (NPA 61.11%; 95% CI, 38.62 – 79.69%), giving an overall concordance of 84.00% (95% CI, 71.49 – 91.66%; **Table 3** & **Supplementary Table 4**). PPA was high (>91%) across the four *EGFR* variant types (exon 19 deletion, L858R, T790M, and uncommon G719X/S768I/exon 20 insertion variants; **Fig 4A**), with 1 EGFR L858R detected by cobas but not LiquidHALLMARK. To resolve the true nucleotide identity of this potential false negative variant, plasma cfDNA from this sample was orthogonally tested by AS-PCR, which corroborated the absence of EGFR L858R. Hence, orthogonal AS-PCR validation coupled with the established 99% specificity of cobas for EGFR L858R alterations (benchmarked against tissue testing) [34] suggest that the potential false negative by LiquidHALLMARK could have been a false positive via cobas testing, supporting a 100% PPA of LiquidHALLMARK with cobas. Across the four *EGFR* variant types, NPA also remained high (>93%; **Fig 4B**), with LiquidHALLMARK detecting seven additional variants across 0.05 – 0.33% VAF, including 2 L858R, 2 exon 19 deletion, 2 uncommon (G719A and S768I), and 1 T790M variants, which are within the range of variants detectable by the cobas assay (**Fig 4C** & **Supplementary Table 4**). To resolve these potential false positive calls, plasma cfDNA from these samples were orthogonally validated by AS-PCR. Of the seven variants, six were positively detected by AS-PCR, including one exon 19 deletion (EGFR E746_A750del, detected at 0.15% VAF), one G719A (detected at 0.08% VAF), and one T790M variant (detected at 0.23% VAF) specifically detected by melt curve amplification below the reportable range of the AS-PCR assay. The remaining variant (EGFR S768I, detected at 0.12% VAF) was not covered by the AS-PCR assay and could not be orthogonally confirmed. These results are consistent with the established 25 – 100 copies/mL plasma [35] and reported 0.1 – 0.8% VAF [36] LoD of the cobas assay, with all discordant variant calls falling below 0.4% VAF (**Fig 4C**), establishing the exceptional sensitivity of LiquidHALLMARK.

**Table 3.** Comparison of LiquidHALLMARK with cobas EGFR Mutation Test v2 for the detection of EGFR L858R, exon 19 deletion, T790M, and G719X/S768I/exon 20 insertion variants.

| LiquidHALLMARK® | Reference assay (cobas® EGFR Mutation Test v2) |  |  |  | PPA: 96.88% (84.26 – 99.84%)<br>NPA: 61.11% (38.62 – 79.69%)<br>OPA: 84.00% (71.49 – 91.66%) |
| --- | --- | --- | --- | --- | --- |
|  |  | Positive | Negative | Total |  |
|  | Positive | 31 | 7 | 38 |  |
|  | Negative | 1 | 11 | 12 |  |
|  | Total | 32 | 18 | 50 |  |

PPA, positive percent agreement; NPA, negative percent agreement; OPA, overall percent agreement.

**Figure 4.**
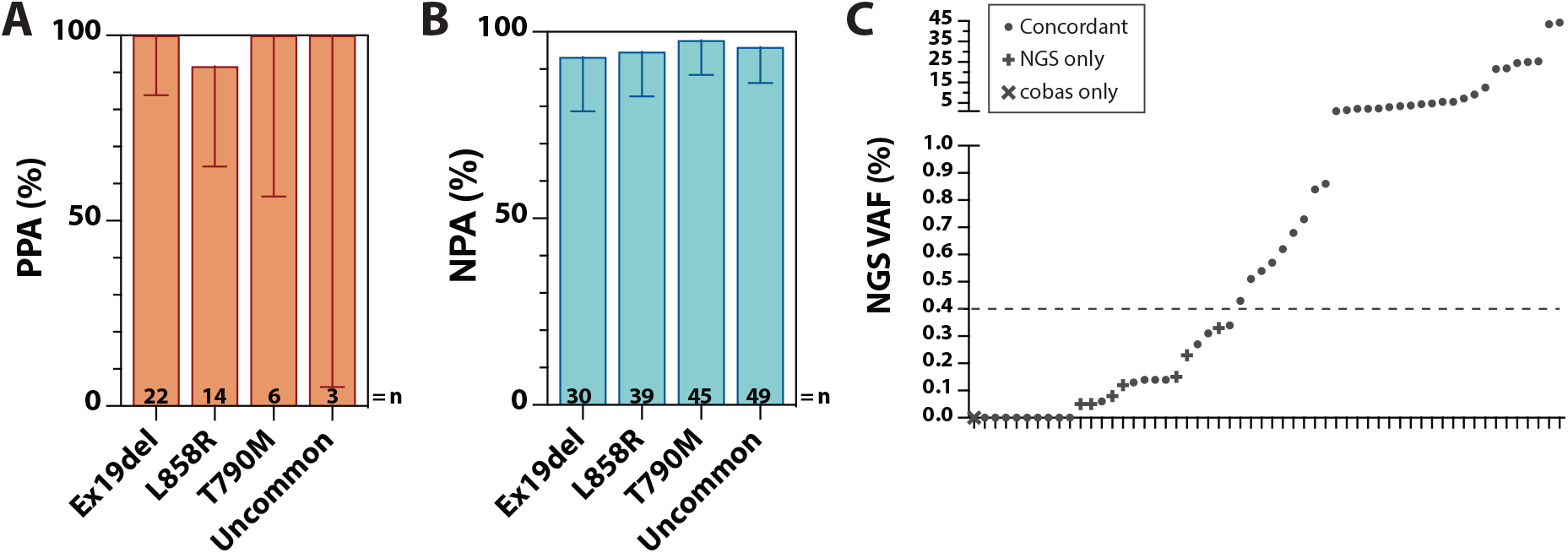
Clinical validation for detection of *EGFR* alterations. **(A**) Positive percent agreement (PPA), and (**B**) negative percent agreement (NPA) of LiquidHALLMARK *EGFR* variant calls relative to cobas EGFR Mutation Test v2 in 50 clinical lung cancer samples. Uncommon *EGFR* variants include G719X, S768I, and exon 20 insertions. Error bars in **A** and **B** represent 95% CI. (**C**) Positive and negative concordance of LiquidHALLMARK *EGFR* variant calls relative to cobas EGFR Mutation Test v2. VAF, variant allele frequency. Each data point represents a negative sample (VAF = 0%) or *EGFR* variant; •, concordant call between LiquidHALLMARK and cobas; x, detected only by cobas; +, detected only by LiquidHALLMARK (NGS). Above 0.4% VAF, both assays were concordant for all detected *EGFR* variants.

### Clinical utility of the LiquidHALLMARK Assay

While validation studies are crucial for examining the technical performance of an assay, insight into the performance of an assay in a real-world setting is equally, if not more, informative on the utility of a liquid biopsy test in guiding clinical decisions. To that end, we examined all samples submitted to our CAP- and CLIA-accredited laboratory to elucidate real-world assay performance metrics. Within the study period from January 2018 to May 2021, a total of 1,592 samples were analyzed and reported (100% report rate), with 52.1% of the clinical volume originating from lung cancer patients (**Fig 5A**). While the assay is intended for advanced cancer patients to aid clinical therapeutic decision making, a significant portion (n = 71; 4.5%) of the clinical volume comprised screening cases in healthy individuals, or suspected cancer cases. Additionally, samples from patients with localized tumors constituted 8.7% of the clinical volume. Overall, 73.6% (1120/1521) of cancer samples harbored at least one detectable genetic variant (ctDNA positive), including 40.6% of samples from localized tumors and 78.5% of samples from metastatic tumors. When non-genetic biomarkers (EBV/HBV/MSI) were included, the detection rate increased to 74.8% (n = 1137). While ctDNA detection was largely uniform across most tumor types, several cancers such as kidney cancer and melanoma displayed lower ctDNA detection rates (**Fig 5A**), consistent with previous reports on tumor type-specific ctDNA prevalence [15]. Among ctDNA-positive samples, a median of two variants were detected per sample, with a median VAF of 0.86% (range 0.01 – 95.30%). Additionally, more than half of ctDNA-positive cancer samples contained at least one variant ≤0.3% VAF; this was again consistent across tumor type, highlighting the ability of the assay to sensitively detect variants at sub-1% VAFs. Across all non-cancer screening samples (n = 61), two likely pathogenic SNV variants (JAK2 V617F, detected at 0.81% VAF, and TP53 M246I, detected at 0.40% VAF) were observed, while no INDELs, CNAs, gene fusions, or non-genomic biomarkers, were detected, giving a per-sample clinical specificity of 96.72%, or per-base specificity of ≥99.9999% (2 SNVs across 3.59 million base pairs, or 61 samples x 58.8 kb).

**Figure 5.**
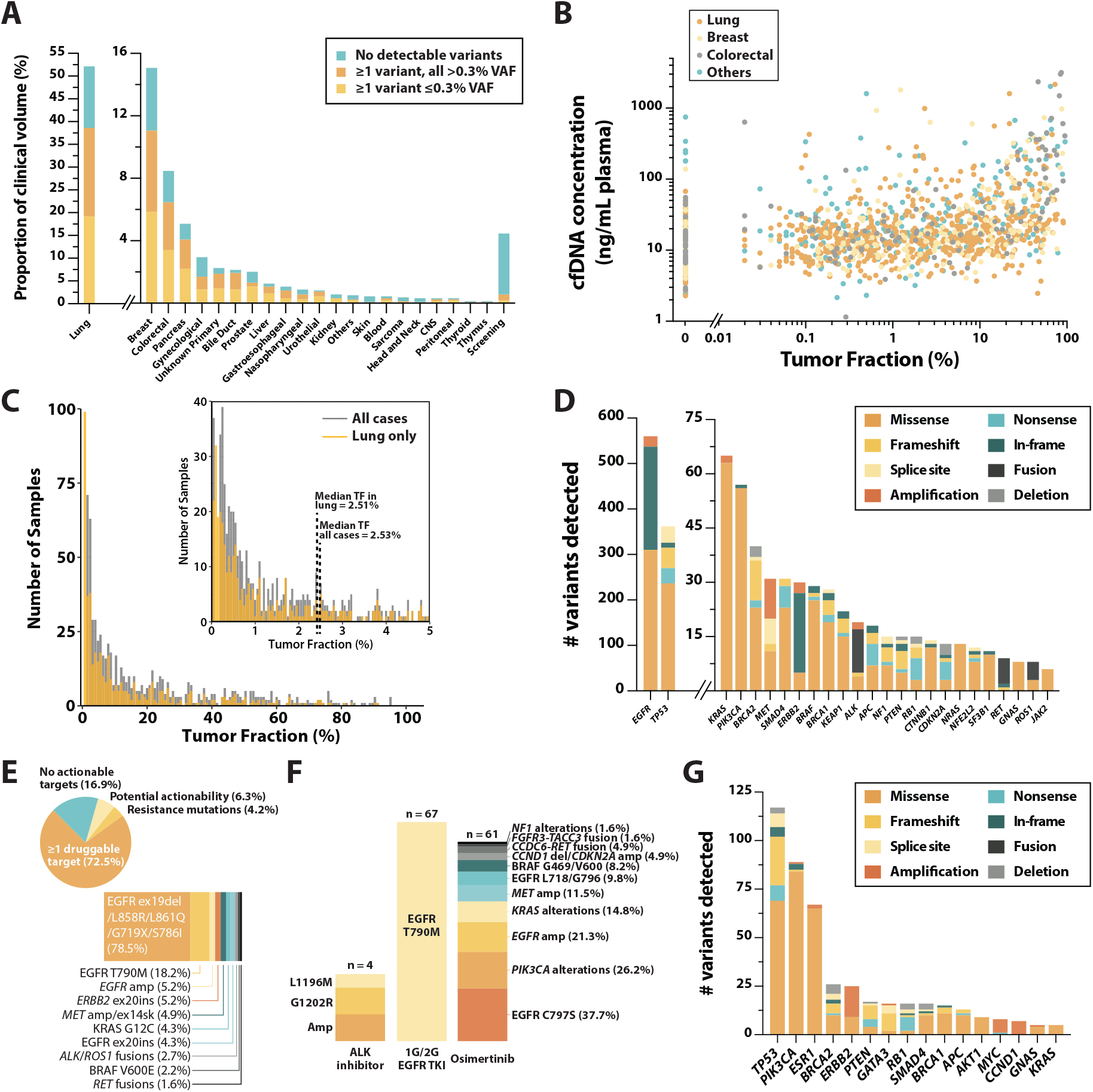
Clinical utility of the LiquidHALLMARK^®^ assay. **(A)** Proportion of clinical volume (n = 1,592) by tumor origin. Samples with no detectable variants are shaded in blue, while positive samples are shaded in yellow if at least 1 variant was detected at a variant allele frequency (VAF) ≤0.3% and orange if all variants detected were >0.3% VAF. CNS, central nervous system. **(B)** Correlation between cell-free DNA (cfDNA) concentration and tumor fraction, approximated as the variant with the highest VAF observed in any given sample. Lung, breast, and colorectal cancer samples are highlighted in orange, yellow and gray respectively; all other tumor types are highlighted in blue. **(C)** Distribution of positive samples by tumor fraction. The distribution for positive lung cancer samples is shown in yellow. **(D)** Prevalence and variant type of the top 25 genes altered in lung cancer samples. Each color represents a different variant type. (**E**, top) Proportion and breakdown of 615 ctDNA-positive lung cancer samples with druggable targets. (**E**, bottom) Prevalence and breakdown of druggable targets in lung cancers with guideline-approved biomarkers. (**F**) Prevalence and breakdown of alterations associated with resistance observed in lung cancer patients treated with an ALK inhibitor, 1^st^ or 2^nd^ generation (1G/2G) EGFR tyrosine kinase inhibitor (TKI), or osimertinib. Ex20ins, exon 20 insertion; ex14sk, exon 14 skipping; amp, amplification; del, deletion. (**G**) Prevalence and variant type of the top 15 genes altered in breast cancer samples. Each color represents a different variant type.

Given that the presence of ctDNA correlates with tumor burden [37–40], we evaluated the correlation between plasma cfDNA concentration and tumor fraction (approximated as the variant with the highest VAF observed in any given sample). Plasma cfDNA concentration was positively correlated with TF, although this varied across different tumor types; for instance, ctDNA-positive colorectal cancer samples possessed a higher median concentration (median 25.57 ng/mL plasma) compared to ctDNA-positive lung cancer samples (median 15.50 ng/mL; **Fig 5B**). While the median concentration of ctDNA-negative samples was lower (10.87 ng/mL, range 2.29 – 746.67 ng/mL) compared to that of ctDNA-positive samples (17.56 ng/mL, range 1.15 – 3160 ng/mL), a significant portion of ctDNA-negative samples possessed high cfDNA concentrations (**Fig 5B**), highlighting a potential limitation of targeted mutation-based liquid biopsy approaches in accurately determining tumor burden. Across all ctDNA-positive samples, the median TF was 2.53% (2.51% in lung cancer samples), with 20.2% of samples having a TF ≤0.3% (19.4% in lung cancer samples; **Fig 5C**), emphasizing the importance of high sensitivity in liquid biopsy assays.

### Clinical actionability

A cornerstone of NGS-based liquid biopsy assays is the ability to detect clinically actionable alterations not easily detected via alternative testing methods. Hence, we further analyzed the prevalence of driver genes and clinically relevant variants detected within the 1,592 clinical samples. In line with known prevalence of significantly mutated genes in cancer [41,42], *TP53, KRAS, PIK3CA, APC, SMAD4*, and *PTEN* were among the most frequently altered genes across all ctDNA-positive samples (**Supplementary Fig 4A**). In addition, *EGFR* alterations were detected in 36.1% of all ctDNA-positive samples; this was driven by the large proportion of lung cancers among clinical samples. Indeed, in ctDNA-positive lung cancer samples, the detection rate of *EGFR* alterations was 62.8%, with more than a third of detected *EGFR* alterations being in-frame exon 19 deletions (**Fig 5D, Supplementary Fig 4B**). Significant alteration frequencies were also observed in a repertoire of clinically relevant genes including *KRAS, PIK3CA, MET, ERBB2, BRAF*, and *ALK*, as well as driver genes with potential actionability such as *NF1, KEAP1*, and *NFE2L2* (**Fig 5D**). Importantly, of the 615 ctDNA-positive lung cancer samples, 83.1% (n = 511) harbored at least one genetic biomarker possessing biological evidence for actionability, including 72.5% (n = 446) with ≥1 biomarker with a guideline-approved drug, and an additional 4.2% (n = 26) and 6.3% (n = 39) harboring either biomarkers predicting drug resistance or with biological evidence suggesting potential clinical actionability (both drug sensitivity and resistance) respectively (**Fig 5E** top). Significantly, 175 of the 511 lung cancer samples with actionable biomarkers (28.5% of all ctDNA-positive lung cancer samples) harbored an actionable biomarker at low VAF (≤0.3%). Sensitizing *EGFR* mutations (EGFR L858R, exon 19 deletion, or L861Q/G719X/S786I) were by far the most prevalent druggable target, being observed in 78.5% of lung cancer samples with guideline-approved biomarkers, while EGFR T790M, exon 20 insertions, and amplifications, were also observed at significant frequencies (**Fig 5E** bottom). Beyond drug sensitivity, the prevalence of alterations that predict treatment resistance was also further analyzed. On-target *ALK* mutations [43] including ALK G1202R, ALK L1196M, and *ALK* amplification were observed in 14.8% (4/27) of ALK inhibitor-treated patients; EGFR T790M [44] was observed in 37.0% (67/181) of first- or second-generation EGFR TKI-treated patients, and a range of resistance mutations (in accordance with literature on osimertinib resistance [45]), including EGFR C797, *PIK3CA* and *KRAS* alterations, *MET* amplification, copy number changes in cell cycles genes, *FGFR3* and *RET* fusions, was observed in 37.4% (61/163) of osimertinib-treated patients (**Fig 5F**). Strikingly, 32.8% (22/67) of all EGFR T790M resistance mutations in first- or second-generation EGFR TKI-treated patients were detected at VAF ≤0.3%, while 42.6% (26/61) of all resistance mutations detected in osimertinib-treated patients were observed at VAF ≤0.3%. In breast cancer, clinically relevant alterations in *PIK3CA, ESR1, BRCA2*, and *ERBB2* were among the most prevalent (**Fig 5G, Supplementary Fig 4C**); 46.8% (59/126) of HR+ HER2-breast cancer samples harbored either an *ESR1* mutation predicting resistance to aromatase inhibitors [46], or a *PIK3CA* mutation predicting sensitivity to PI3K pathway inhibition by alpelisib [47], with 28.8% (17/59) of these samples harboring concurrent *PIK3CA* or *ESR1* mutations (**Supplementary Figure 5A**). Additionally, loss-of-function alterations in *RB1*, predicted to contribute to CDK4/6 inhibitor resistance [48], were observed in 13.3% (6/56) of CDK4/6 inhibitor-treated HR+ HER2-breast cancer patients. In colorectal cancer patients, 54.1% of all samples harbored a mutation known to confer anti-EGFR antibody resistance [49,50] (**Supplementary Figure 5B**). Across the three cancer types, actionable variants were among the most frequently observed variants, with VAFs ranging from 0.01 – 94.30% (**Table 4**). Collectively, these results demonstrate the capability of LiquidHALLMARK to guide clinical therapeutic decisions.

**Table 4.** Median variant allele fraction (VAF) of top 10 mutated genes in clinical lung, breast, and colorectal samples.

| Gene | Variant | Variant Type | Median VAF % (Range) |
| --- | --- | --- | --- |
| **Lung cancer, n = 615** |  |  |  |
| EGFR | Ex19del | INDEL | 2.99% (0.01 – 90.40%) |
| EGFR | L858R | SNV | 2.42% (0.01 – 73.20%) |
| EGFR | T790M | SNV | 0.72% (0.04 – 67.03%) |
| EGFR | C797S | SNV | 1.50% (0.14 – 63.90%) |
| EGFR | Amplification | CNA | 2.06X (1.42 – 8.51X) |
| KRAS | G12C | SNV | 3.80% (0.06 – 26.80%) |
| PIK3CA | E545K | SNV | 2.90% (0.03 – 48.58%) |
| ERBB2 | Y772_A775del | INDEL | 0.84% (0.09 – 23.67%) |
| KRAS | G12D | SNV | 1.49% (0.02 – 59.94%) |
| MET | Amplification | CNA | 2.50X (1.64 – 6.73X) |
| ALK | EML4-ALK | Fusion | 3.20% (0.19 – 54.20%) |
| **Breast cancer, n = 176** |  |  |  |
| PIK3CA | H1047R | SNV | 3.09% (0.06 – 61.90%) |
| ESR1 | D538G | SNV | 1.19% (0.11 – 68.06%) |
| ERBB2 | Amplification | CNA | 2.95X (1.40 – 10.09X) |
| PIK3CA | E545K | SNV | 7.60% (0.22 – 41.19%) |
| PIK3CA | E542K | SNV | 2.30% (0.11 – 69.50%) |
| AKT1 | E17K | SNV | 2.80% (0.25 – 23.11%) |
| ESR1 | Y537S | SNV | 1.46% (0.13 – 32.70%) |
| TP53 | Y163S | SNV | 0.36% (0.26 – 0.68%) |
| CCND1 | Amplification | CNA | 2.30X (2.01 – 3.99X) |
| MYC | Amplification | CNA | 2.42X (1.66 – 5.41X) |
| ESR1 | E380Q | SNV | 2.60% (0.06 – 21.22%) |
| **Colorectal cancer, n = 103** |  |  |  |
| KRAS | G12D | SNV | 15.90% (0.06 – 76.30%) |
| KRAS | G13D | SNV | 0.37% (0.06 – 27.12%) |
| KRAS | G12V | SNV | 0.58% (0.02 – 34.81%) |
| KRAS | Q61H | SNV | 3.91% (0.23 – 34.50%) |
| MYC | Amplification | CNA | 2.00X (1.55 – 23.31X) |
| APC | S1465Wfs*3 | INDEL | 0.61% (0.27 – 48.41%) |
| TP53 | R273H | SNV | 4.35% (0.35 – 86.10%) |
| APC | c.835-8A>G | SNV | 1.18% (0.66 – 52.16%) |
| APC | R1450* | SNV | 12.82% (0.45 – 40.30%) |
| APC | R876* | SNV | 2.06% (0.92 – 48.99%) |
| BRAF | V600E | SNV | 6.76% (0.02 – 38.90%) |
| PIK3CA | E542K | SNV | 9.05% (0.23 – 20.67%) |
| PTEN | R130* | SNV | 0.17% (0.08 – 21.40%) |
| TP53 | G245S | SNV | 28.01% (0.99 – 61.59%) |
| TP53 | Y220C | SNV | 84.40% (59.89 – 94.90%) |

### Molecular profiling to delineate origins of cancers of unknown primary

Broad genomic coverage also aids the molecular profiling and differential diagnosis of cancers of unknown primary (CUP). Of the 36 CUP cases analyzed, 80 unique genetic alterations across 34 genes were detected in 30 cases, giving a ctDNA detection rate of 83.3% (**Supplementary Fig 6**). Detection of tumor type-specific alterations, including *AR, ESR1, GNAS*, and *EGFR* alterations in seven samples, as well as EBV, HBV, and MSI positivity in three samples, informed potential tumor origins. Within genes with high prevalence of mutations, tumor specific prevalence biases based on the COSMIC database [51] could be used to discriminate between tumor types. For instance, several *TP53* alterations identified favored a large intestinal origin (R273C, R273H), while others favored a lung origin (K305^⋆^, V157F, R110P).

### Cancer monitoring through alternative non-genomic biomarkers

In tumor types where the prevalence of conventional genetic biomarkers is low, such as nasopharyngeal cancers (NPC), viral load quantification acts as a useful marker of tumor burden [52]. Of the NPC samples analyzed, 64.3% (9/14) were ctDNA positive, with *TP53* alterations being the most prevalent. By contrast, 69.2% (9/13; one sample was not tested for EBV) of NPC samples were EBV positive (**Supplementary Fig 7A)**, including two ctDNA-negative samples, enabling an additional diagnostic yield of 14.3%. In liver cancer, although 85.0% (17/20) of samples were ctDNA positive and only 55.6% (10/18; 2 samples were not tested for HBV) of samples were positive for HBV (**Supplementary Fig 7B**), one ctDNA-negative sample was HBV positive, providing an additional diagnostic yield of 5.0%. The detection of microsatellite instability also contributed to diagnostic yield; 16 samples were identified as MSI positive, including six MSI-high samples and 10 MSI-low samples. Of these, three were negative for any ctDNA-based genetic variant.

## Discussion

Tissue biopsy has historically been the gold standard of cancer diagnostics, enabling the histopathological analysis of tumors, and since the advent of precision oncology, facilitates the molecular profiling of tumors to inform therapeutic decisions [53]. Yet, as the field of molecular biology advances, the limitation of tissue biopsy for molecular profiling becomes increasingly evident. Critically, the isolated snapshot provided by a single tissue biopsy is unable to wholly recapitulate a tumor, particularly given extensive evidence of intratumoral heterogeneity, which may impact therapeutic options [54–56]. This emphasizes the need for serial biopsies, an option that is often infeasible given the invasive nature of tissue biopsies and their accompanying clinical risks, potential surgical complications, and economic costs [57]. In scenarios where tumor accessibility is an issue, obtaining even a single tissue biopsy can be onerous [53]. In light of this, liquid biopsy presents a minimally invasive opportunity to interrogate tumor biology without additional risks to patients, enabling early cancer detection, prognostication, and disease monitoring through a simple blood draw. Several cfDNA-based liquid biopsy companion diagnostics currently exist; these assays probe specific molecular targets to predict drug sensitivity, for instance, *EGFR* in the cobas EGFR Mutation Test v2 and *PIK3CA* in the therascreen PIK3CA RGQ PCR Kit [10]. As the rapidly expanding repository of targeted therapies grows, the need for more comprehensive panels that probe a wider spectrum of vulnerabilities becomes increasingly imperative.

Here, we present the analytical and clinical validation of LiquidHALLMARK, an amplicon-based NGS liquid biopsy assay which interrogates 80 cancer-related genes for SNVs, INDELs, CNAs, and gene fusions, as well as additional biomarkers including oncogenic viruses (EBV and HBV) and MSI. These results underscore the exceptional sensitivity (LoD_90_ of 0.1% for SNVs and INDELs, and 0.5% for fusions), specificity (99.98% at 5% and 1% VAF, and 99.11% at 0.1% VAF for SNVs and INDELs), precision, and accuracy of the LiquidHALLMARK panel. Across 674 observations in 49 unique alterations, the assay demonstrated highly quantitative and reproducible VAF measurements. Orthogonal validation of LiquidHALLMARK *EGFR* variant calls with a reference AS-PCR method in 355 lung cancer specimens revealed an overall concordance of 93.80% (PPA 95.49%; NPA 92.79%), highlighting the excellent accuracy of the assay. Comparison of LiquidHALLMARK against the FDA-approved cobas EGFR Mutation Test v2 for 50 lung cancer specimens demonstrated an overall concordance of 84.00% (PPA 96.88%; NPA 61.11%), with eight discordant calls (LiquidHALLMARK VAF 0.05 – 0.33%) between LiquidHALLMARK and cobas. Of these, seven were resolved by orthogonal AS-PCR testing, supporting the calls made by LiquidHALLMARK. These results exemplify the high sensitivity of LiquidHALLMARK, even at VAFs as low as 0.05%. In addition, the assay exhibits uniformly high sequencing depth across the targeted regions of genome coverage, recovering a median 69.6% of unique input DNA molecules (range 42.0 – 98.4%), with 98.7% of calls having a unique coverage of >1000X. Several additional capabilities of the assay are highlighted in these validation studies. The ability of the platform technology to detect novel fusions was exemplified by an overall concordance of 100% against orthogonal testing methods for structural rearrangements in PD-L1 that disrupt its 3’-UTR [29]. Further, the quantitative detection of oncogenic viruses such as EBV and HBV demonstrated excellent linearity (R^2^ >0.95) across six orders of magnitude, down to an LoD of 2 IU/mL plasma.

When used within a real-world setting for 1,592 clinical samples, 74.8% of cancer samples harbored at least one clinically useful biomarker for drug targeting, clinical trial inclusion, or disease monitoring, including 36.2% with on-label drug recommendations, and an additional 5.7% and 8.1% harboring biomarkers associated with treatment resistance and potential actionability respectively. Importantly, the diverse spectrum of alterations across the 80 cancer-related genes covered in the panel is highlighted by the 1607 unique somatic variants identified across all clinical samples, emphasizing the necessity of broad genomic coverage. Underpinning the high sensitivity of the assay is the frequent detection of samples with TFs ≤ 0.3%, with variants routinely detected below the LoD; indeed, 10.2% of all reported variants were found at VAFs between 0.01% and 0.09%, with calls being supported by high variant allele read counts and a high depth of coverage. In lung cancer, the assay identified an actionable biomarker in 83.1% of ctDNA-positive cases, including 28.5% of cases where the actionable biomarker was detected at a low VAF of ≤0.3%. Significantly, nearly a third of first- or second-generation EGFR TKI-associated EGFR T790M resistance mutations, and more than two-fifths of osimertinib-associated resistance mutations, were detected at low VAFs ≤0.3%. These findings reinforce the necessity of high sensitivity in liquid biopsies, particularly in lung cancers where such actionable findings can directly impact treatment outcomes in patients. Besides the sensitive detection of SNVs and INDELs, gene fusions in *ALK, RET*, or *ROS1* were detected in 2.9% of ctDNA-positive lung cancer cases, exemplifying the clinical applicability of an amplicon-based assay for the detection of novel fusions. The specificity of the assay, enhanced through error suppression via UMI sequencing coupled with statistical background error-modelling, was demonstrated through the real-world analysis of non-cancer screening patients. Across 61 non-cancer samples, two likely pathogenic variants, JAK2 V617F and TP53 M246I, were detected at 0.81% and 0.40% VAF respectively, demonstrating a per-sample specificity of 96.72% or per-base specificity of ≥99.9999%. Furthermore, given that JAK2 V617F and mutations in *TP53* are two of the most common somatic alterations detected in plasma cfDNA due to clonal hematopoiesis (CH) [29], the possibility that these two mutations arose due to CH cannot be ruled out. Taken together, these results underline the excellent sensitivity and specificity of the LiquidHALLMARK assay within a real-world clinical setting. In addition, the inclusion of less commonly-targeted oncogenic biomarkers in LiquidHALLMARK enhances diagnostic yield. For instance, EBV clearance in ctDNA has been investigated as a surrogate for clinical outcomes in the only prospective, randomized, interventional study in the minimal residual disease (MRD) setting [58] and an EBV qPCR assay has also been used as a blood-based primary screening test for nasopharyngeal cancer [59], while MSI is an actionable biomarker that is routinely tested in tumor tissue but is currently limited in liquid biopsy assays.

EBV and HBV detection enabled a further diagnostic yield of 14.3% and 5.0% respectively in nasopharyngeal and liver cancers, while MSI detection provided on-label drug recommendations in six samples and additional diagnostic yield in three ctDNA-negative samples.

Collectively, these results establish the performance of an amplicon-based liquid biopsy assay in a niche where the utility of hybrid-capture methods has been much more extensively evaluated [15,24]. Although conventional whole-exome amplicon-based NGS sequencing has been associated with lower uniformity of depth [60], this limitation is easily circumvented in targeted liquid biopsy panels by rational design, particularly given that cfDNA is fragmented in a non-random fashion [61–63]. Indeed, LiquidHALLMARK demonstrates highly uniform sequencing depth, with an average 97.1% of calls having a depth no less than 0.2X of the mean panel coverage and 79.0% of calls having no less than half the mean panel coverage. This, coupled with the unmatched specificity of PCR combined with UMI and statistical error-modelling, affords the LiquidHALLMARK assay exquisite sensitivity, even at low cfDNA input amounts of 20 ng (containing approximately 6000 haploid genome equivalents), ensuring a high assay robustness (100% report rate in 1,592 consecutive clinical samples) and reliability of test results, and highlighting the value and utility of amplicon-based methods in monitoring oncology treatment sensitivity and resistance. Additional studies to demonstrate clinical validity and utility are ongoing.

## Materials and methods

### Sample collection and processing

Two 10 mL tubes of whole blood were collected for each patient in Streck Cell-Free DNA BCT (Streck), shipped to Lucence Diagnostics at ambient temperature, and processed within 6 – 48 h of sample collection. Upon sample receipt, samples were accessioned and both 10 mL tubes of whole blood per patient were fractionated to isolate plasma via centrifugation at 1,600 *g* for 10 min at 4°C. Isolated plasma was clarified via further centrifugation at 16,000 *g* for 10 min at 4°C before being aliquoted and stored at -20°C or processed immediately for cfDNA extraction.

### cfDNA extraction, library preparation, and sequencing

Cell-free DNA (cfDNA) extraction, library preparation, and sequencing were performed in a CLIA-certified, CAP-accredited laboratory (Lucence Diagnostics). Briefly, cfDNA was extracted from 4 – 8 mL plasma (from one tube of whole blood) using the QIAamp Circulating Nucleic Acid Kit (Qiagen) and quantified on a Qubit® 2.0 fluorometer (Thermo Fisher), yielding a median of 14.13 ng cfDNA/mL plasma (range 1.15 – 3160 ng/mL plasma). For each sample, 10 – 20 ng of extracted cfDNA was converted to an Illumina-compatible sequencing amplicon library, first via limited PCR amplification using random oligonucleotide barcodes (IDT), then enriched with sample-specific barcodes and Illumina sequencing adapters. For the detection of fusions, 7.5 – 15 ng of extracted cfDNA was barcoded via limited PCR amplification before undergoing A-tailing and single-ended ligation of partial Illumina sequencing adapters, followed by enrichment with sample-specific barcodes and Illumina sequencing adapters. Libraries were purified using AMPure XP beads (Beckman Coulter) and quantified using the KAPA Library Quantification Kit (Roche) before being pooled for sequencing by synthesis on a NextSeq 550 (Illumina) to generate paired-end reads (300 cycles).

### Bioinformatics processing and variant calling

Illumina binary base call sequencing files were demultiplexed and converted to fastq file format using the bcl2fastq v2.20 software (Illumina) before being processed with an in-house bioinformatics pipeline (Lucence Diagnostics) for base quality filtering (Phred quality score <15), adapter trimming, and consensus UMI clustering. Consensus reads were then aligned to the hg19 reference genome using BWA-MEM v0.7.17 [64]. For SNV and INDEL detection, consensus reads were compared against position- and sequence run-specific noise level profiles trained using a curated selection of 110 past samples run through the in-house pipeline to estimate the probability of noise for each variant in a sample. Position-specific *p*- value cutoffs were used to discriminate between true and noise variants. All true variants found within coding sequences and splice sites, as well as pathogenic variants within promoter regions based on ClinVar classification were reported [65]. For fusion detection, paired-end reads were mapped to the reference genome and alignment information including directionality and soft-clipped sequences were analyzed to identify potential fusion events. To detect CNAs, probe-level consensus coverage was self-normalized to a set of invariant genomic positions and compared against the normalized coverage distribution constructed from baseline samples before being aggregated to the gene level (z-score) [66]. MSI status was determined by comparing deletion lengths at six repetitive loci (NR27, NR24, BAT25, MONO27, BAT26, and NR21) against a reference baseline; samples were considered MSI-low if one of six markers deviated from the baseline and MSI-high if two or more markers deviated from the baseline. Viral loads of HBV and EBV were quantified based on consensus read coverage using a standard curve method, generated using NIBSC EBV and HBV standards. Alterations were categorized as having strong biological evidence for actionability if the therapeutic indications were FDA-approved, recommended by NCCN or other professional guidelines, or supported by results from well-powered phase III clinical trials with expert consensus. Alterations with actionability evidence in multiple small published studies or case reports were categorized as having potential clinical actionability.

### Contrived sample characteristics

#### SNVs and INDELs

To determine SNV and INDEL detection performance, the reference standard sets HD780 and Tru-Q, as well as the HD786 reference standard, were used (Horizon Discovery). The HD780 Multiplex I cfDNA Reference Standard Set contains cell-line-derived DNA fragmented to an average of 160 bp, and harbors eight variants, including six SNVs and two INDELs, in four genes (*EGFR, KRAS, NRAS*, and *PIK3CA*) at 5%, 1%, and 0.1%, and 0%

VAFs. The Tru-Q Reference Standard set contains six standards, covering 38 SNVs and two INDELs across 19 genes at 5%, 1%, and 0% VAF, as well as four additional endogenous variants at VAF ≥8%. To generate the Tru-Q 0.1% VAF standard, Tru-Q7 (1% VAF standard) was diluted tenfold in Tru-Q0 (wild-type standard). The HD786 Structural Multiplex cfDNA Reference Standard contains six SNVs at 5 – 5.6% VAF, four INDELs at 5.3 – 5.6% VAF, one INDEL at 2.5% VAF, and two endogenous SNVs ≥16% VAF. All alterations are verified by ddPCR. A full list of alterations covered by the three reference standards is found in **Supplementary Table 5**.

#### Fusions

In addition to the SNVs and INDELs described above, the HD786 reference standard also harbors a *SLC34A1-ROS1* fusion at 5.6% and *CCDC6-RET* fusion at 5.0%. To generate additional fusion-positive materials, DNA from the *TMPRSS2-ERG* fusion heterozygous cell-line VCaP, the HD664 EML4-ALK Reference Standard, 50% (Horizon Discovery), and the HapMap cell-line NA24129 were fragmented to 200 bp by acoustic shearing on an ME220 Focused-ultrasonicator (Covaris). Admixtures of the three cell-lines in 1:1:8, 1:1:48, and 1:1:98 ratios were performed to generate *TMPRSS2-ERG* and *EML4-ALK* fusion at 5%, 1%, and 0.5% VAF.

#### CNAs

For CNA detection performance, cell-line DNA from MCF7 (Lot #60540387), HCC2218 (Lot #64097142), and HCC827 (Lot #64216187; all obtained from ATCC) and the HapMap cell-line NA24143 were used following acoustic shearing to 200 bp on an ME220 Focused-ultrasonicator. The HCC827 cell-line harbors a 32.6-fold increase in *EGFR* copy number over the diploid number, as verified by the manufacturers via ddPCR. Fragmented HCC2218 DNA was mixed with fragmented NA24143 DNA to generate cell-line DNA admixtures containing *EGFR* copy numbers of 30, 20, 10, 4, 3, and 2.4. The HCC2218 cell-line harbors a 16.32-fold increase in *ERBB2* (HER2) copy number [30], and the MCF7 cell-line harbors an estimated 4.46-fold increase in *NRAS* copy number, 3.34-fold increase in *MYC* copy number, and a 1.80-fold increase in *CDK6* copy number [31,32]. Fragmented MCF7 DNA was mixed with fragmented NA24143 DNA to generate cell-line DNA admixtures containing 4.3 copies of *NRAS* and 3.6 copies of *MYC*, and 3.2 copies of *NRAS* and 2.8 copies of *MYC*. The HD786 reference standard harbors focal *MET* and *MYC* amplification at 4.5 and 9.5 copies respectively.

#### Oncogenic viruses

For EBV and HBV detection, EBV reference standard (1^st^ WHO International Standard for Epstein-Barr Virus, NIBSC code: 09/260) and HBV reference standard (4th WHO International Standard for HBV DNA for NAT, NIBSC code: 10/266) were each spiked into 1 mL plasma at viral loads spanning 0 to 1,000,000 IU before cfDNA was extracted as previously described.

#### MSI

For MSI detection, admixtures of the MSI-positive fragmented (200 bp) RKO cell-line DNA with plasma cfDNA in 1:99, 1:95, and 1:1 ratios were performed to generate MSI-positive DNA at 1%, 5%, and 50% TF.

### Limit of detection

The lower limit of detection is defined as the lowest VAF at which an alteration can be consistently measured ≥90% of the time (LoD_90_), and was determined empirically based on all assay replicates run on all reference standard materials (HD780, HD786, Tru-Q, and cell-line DNA admixtures) used for analytical validation. The LoD was determined as VAFs for SNVs, INDELs, and fusions, copy number fold-changes for CNAs, and IU/mL plasma for EBV and HBV viral loads.

### Per-base specificity

Analytical specificity for SNVs and INDELs was assessed across all base positions interrogated by the panel using all replicates of the reference standard materials HD780, HD786, and Tru-Q. Endogenous SNPs in the parental cell-lines verified by exome sequencing (Horizon Discovery) were excluded from the analyses. Per-base specificity was determined for all variants detected by NGS prior to noise filtering.

### Clinical sample characteristics

Clinical samples used for validation were collected at the Concord Hospital, National University Hospital, and National Cancer Centre in Singapore. Investigations were carried out according to the principles expressed in the Declaration of Helsinki. The 1,592 consecutive clinical samples included in the observational analyses were received as routine clinical samples and underwent standard processing and reporting at a CAP- and CLIA-accredited laboratory (Lucence Diagnostics). Samples collected between January 2018 and May 2021 were used; all patients provided written informed consent and all data was de-identified. These data span three versions of the LiquidHALLMARK assay (80-gene, 61-gene, and 51-gene panels), with the inclusion of a greater number of targeted genes and genomic regions without changing the underlying molecular principle of the assay. SNVs and INDELs were analyzed in 49 – 77 genes, CNAs were analyzed in 12 – 18 genes, and structural rearrangements were analyzed in 3 – 10 genes, depending on the assay version used (see **Supplementary Fig 1**).

### Clinical and orthogonal validation

The reference AS-PCR method [28] used for orthogonal *EGFR* variant detection was performed in-house using variant-specific PCR primer pairs with wild-type- or mutant-specific fluorescent probes for 10 pre-determined *EGFR* variants. The limit of detection (LoD) for the reference AS-PCR method is 0.05% VAF for *EGFR* exon 19 deletions (E746_A750del, L747_P753delinsS, L747_A750delinsP, and L747_T751del) and L861Q, 0.1% VAF for EGFR G719A, G719S, T790M, and L858R, and 0.5% for EGFR G719C. For external validation of *EGFR* variants, 2 mL of plasma cfDNA from deidentified clinical samples were submitted to the Mayo Clinic Laboratories (Rochester, MN) for cobas EGFR Mutation Test v2. Orthogonal EBV detection was performed by PCR as previously described [33].

Contingency tables were constructed to calculate positive percent agreement (PPA), negative percent agreement (NPA), and overall concordance using standard methods [67]; 95% confidence intervals (95% CI) were calculated using a hybrid Wilson/Brown method [68].

## Data Availability

-

