## Supplementary Data for "Analytical and Clinical Validation of an Amplicon-based Next Generation Sequencing Assay for Ultrasensitive Detection of Circulating Tumor DNA"

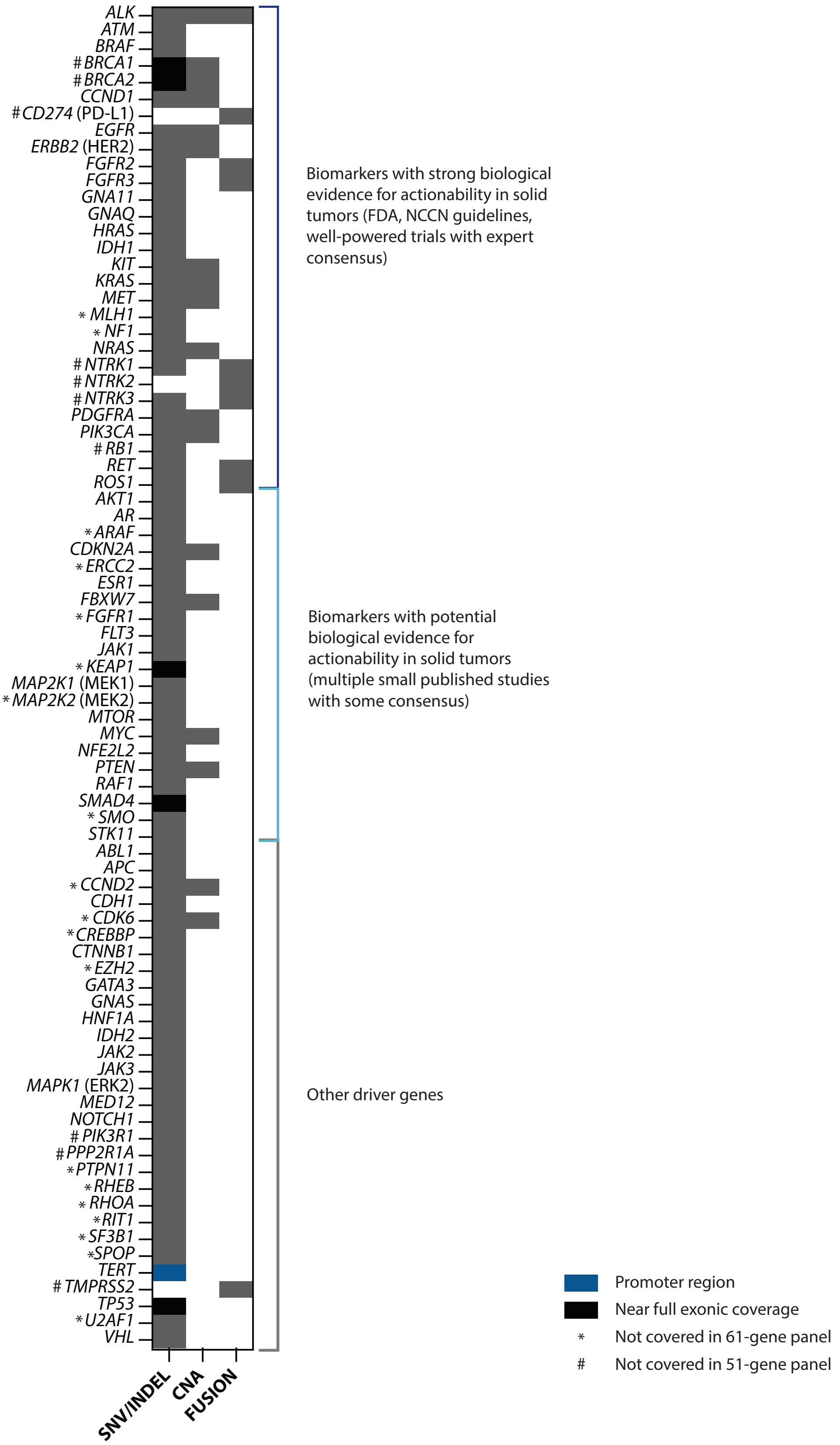


**Supplementary Figure 1 Genome coverage of the LiquidHALLMARK assay.** Genes highlighted in black have near full (>98%) exonic coverage. *TERT* (highlighted in blue) is targeted only in the promoter region. SNV, single nucleotide variant; INDEL insertion/deletion; CNA, copy number alteration. Genes not covered in the 61-gene and 51-gene panels are indicated by * and # respectively.

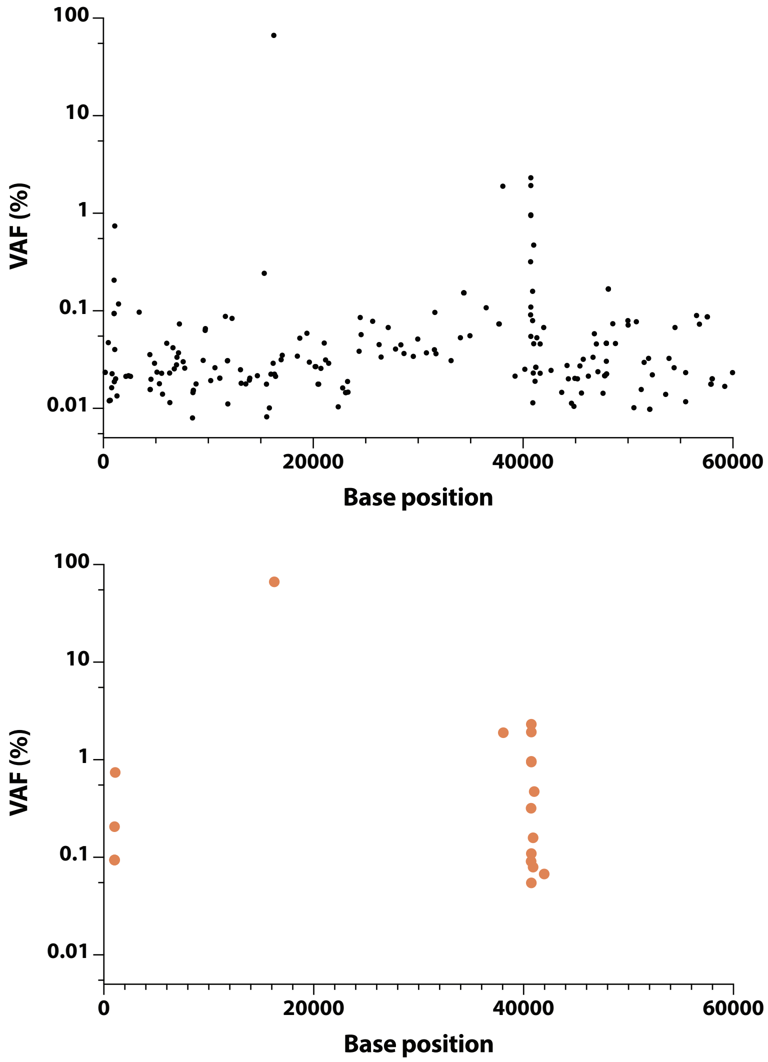


**Supplementary Figure 2 Statistical background error-modelling enables further error suppression of UMI-corrected sequencing data.** Output of all variants (excluding known SNPs) detected in cfDNA following sequencing (top) before and (bottom) after noise filtering based on statistical error-modelling. In this actual sample, a total of 209 variants are detected even with UMI-based error suppression, owing to the ultra-high depth of sequencing. Following noise filtering, only 19 true variants confidently detected above background noise remain.

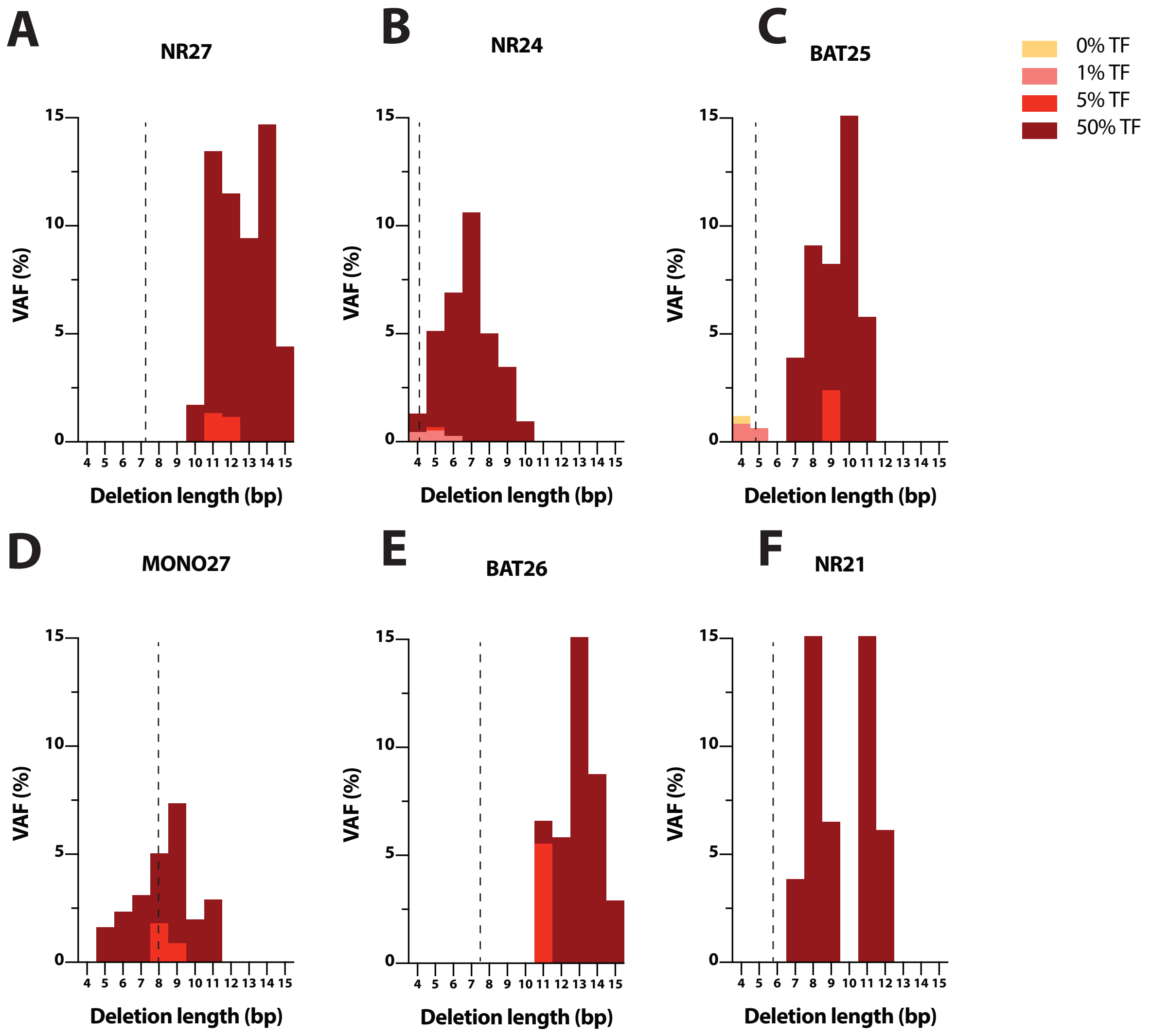


**Supplementary Figure 3 Validation of MSI detection**. Observed deletion length at the (**A**) NR27, (**B**) NR24, (**C**) BAT25, (**D**) MONO27, (**E**) BAT26, and (**F**) NR21 locus in plasma cfDNA (0%) and 1%, 5%, and 50% admixtures of MSI-positive RKO cell-line DNA in plasma cfDNA, representing the tumor fraction (% TF). VAF, variant allele frequency. Broken lines represent the 95$^{th}$ percentile of deletion length in 91 MSI-negative plasma cfDNA and 29 MSI-negative FFPE tissue samples.

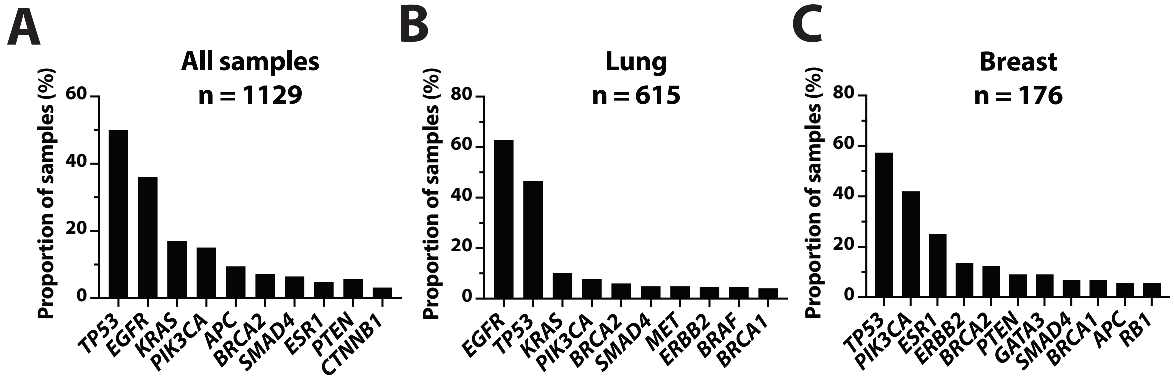


**Supplementary Figure 4 Prevalence of the top 10 genes altered** in (**A**) all ctDNA-positive samples, (**B**) ctDNA-positive lung cancers, and (**C**) ctDNA-positive breast cancers.

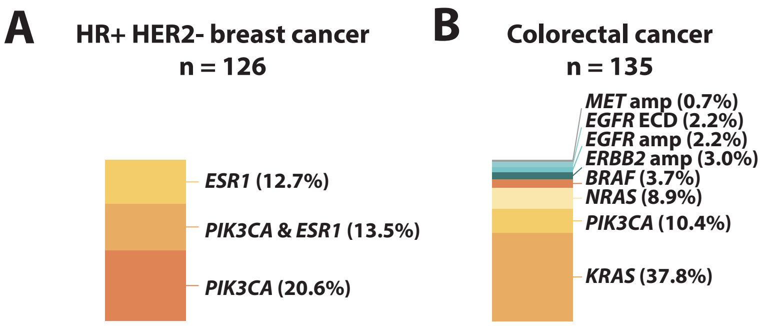


**Supplementary Figure 5 Prevalence of resistance-associated alterations in (A) HR+ HER2- breast cancer and (B) colorectal cancer.** Amp, amplification; *EGFR* ECD, *EGFR* extracellular domain.

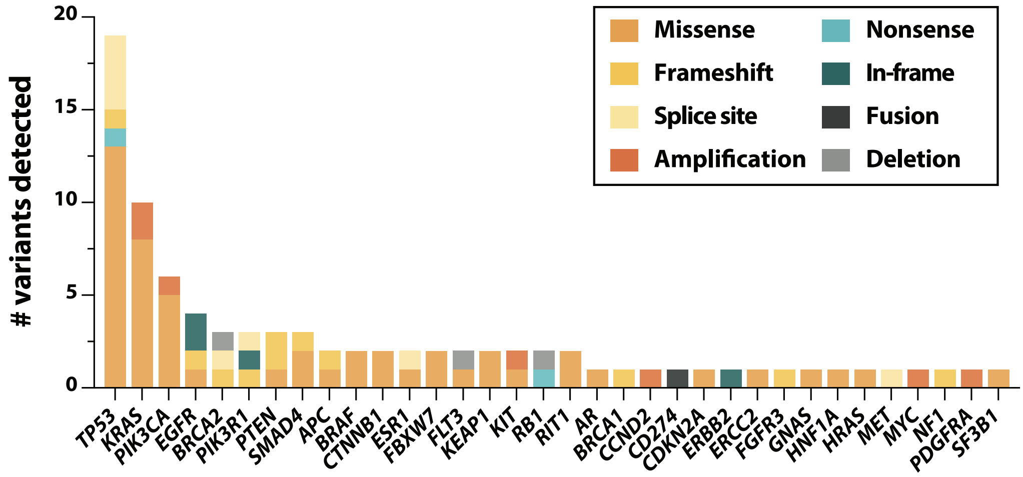


**Supplementary Figure 6 Prevalence and variant type of all alterations detected in cancers of unknown primary (CUP).** Each color represents a different variant type.

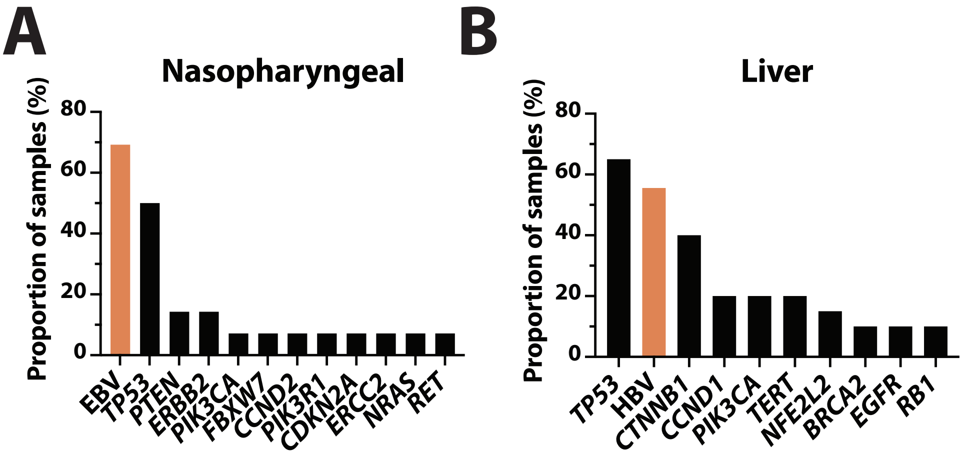


**Supplementary Figure 7 Prevalence of biomarkers altered** in (**A**) nasopharyngeal and (**B**) liver cancer samples. Epstein-Barr virus (EBV) and Hepatitis B virus (HBV) are highlighted in red.

**Supplementary Table 1 Comparison of TissueHALLMARK with a reference assay for the detection of PD-L1 structural rearrangements.**

| | | Reference assay (WGS, targeted, or Sanger Sequencing) | | | |
|---|---|---|---|---|---|
| TissueHALLMARK® | | **Positive** | **Negative** | **Total** | **PPA: 100% (70.09 – 100%)<br>NPA: 100% (83.89 – 100%)<br>OPA: 100% (88.30 – 100%)** |
| | **Positive** | 9 | 0 | 9 | |
| | **Negative** | 0 | 20 | 20 | |
| | **Total** | 9 | 20 | 29 | |

PPA, positive percent agreement; NPA, negative percent agreement; OPA, overall percent agreement.

**Supplementary Table 2 Concordance between cfDNA and matched tissue testing for select clinically relevant mutations and other biomarkers.**

| Alteration/Biomarker | n | PPA (95% CI) | NPA (95% CI) | Tissue testing method |
|---|---|---|---|---|
| *ERBB2* amplification | 9 | 66.67% (35.42 – 87.94%) | - | FISH |
| Microsatellite Instability | 5 | 80.00% (37.55 – 98.97%) | 100% (88.97 – 100%) | Fragment size analysis |

PPA, positive percent agreement; NPA, negative percent agreement.

**Supplementary Table 3 Comparison of LiquidHALLMARK with a reference assay for the detection of EBV.**

| | | Reference assay (EBV BamHI PCR) | | | |
|---|---|---|---|---|---|
| LiquidHALLMARK® | | **Positive** | **Negative** | **Total** | **PPA: 94.12% (73.02 – 99.70%)<br>NPA: 50.00% (2.57 – 97.44%)<br>OPA: 89.47% (68.61 – 98.13%)** |
| | **Positive** | 16 | 1 | 17 | |
| | **Negative** | 1 | 1 | 2 | |
| | **Total** | 17 | 2 | 19 | |

PPA, positive percent agreement; NPA, negative percent agreement; OPA, overall percent agreement.
